# Effects of case- and population-based COVID-19 interventions in Taiwan

**DOI:** 10.1101/2020.08.17.20176255

**Authors:** Ta-Chou Ng, Hao-Yuan Cheng, Hsiao-Han Chang, Cheng-Chieh Liu, Chih-Chi Yang, Shu-Wan Jian, Ding-Ping Liu, Ted Cohen, Hsien-Ho Lin

## Abstract

In the first wave of the COVID-19 pandemic, broad usage of non-pharmaceutical interventions played a crucial role in controlling epidemics^1–6^. However, the substantial economic and societal costs of continuous use of border controls, travel restrictions, and physical distancing measures suggest that these measures may not be sustainable and that policymakers have to seek strategies to lift the restrictions. Taiwan was one of the few countries that demonstrated initial success in eliminating the COVID-19 outbreak without strict lockdown or school closure. To understand the key contributors to the successful control, we applied a stochastic branching model to empirical case data to evaluate and compare the effectiveness of more targeted case-based (including contact tracing and quarantine) and less targeted population-based interventions (including social distancing and face mask use) in Taiwan. We found that case-based interventions alone would not be sufficient to contain the epidemic, even in a setting where a highly efficient contact tracing program was in place. The voluntary population-based interventions have reduced the reproduction numbers by more than 60% and have likely played a critical role at the early stage of the outbreak. Our analysis of Taiwan’s success highlights that coordinated efforts from both the government and the citizens are indispensable in the fight against COVID-19 pandemic.

## Introduction

While the coronavirus disease 2019 (COVID-19) outbreaks currently show no signs of deceleration in the Americas, Africa, and Southeast Asia^7^, some countries successfully contained the first wave of the outbreak with the use of strong interventions such as strict lockdowns and border closures^2–5,8^. Taiwan was initially considered a high-risk country for the COVID-19 outbreak, given its close relationship with China economically and geographically. Nevertheless, six months after the outbreak, Taiwan has one of the lowest per capita incidence and mortality rates of COVID-19 in the world, and no local cases were confirmed between early April and mid-August^9^. Notably, the containment of COVID-19 in Taiwan was achieved without strict lockdown or school closure.^10^

To prevent the healthcare system from being overwhelmed, Taiwan implemented the “containment-as-mitigation” or elimination strategy during the early phase^8,11^. This approach included border control, case-based interventions targeting COVID-19 patients, and population-based measures targeting the general public^11,12^. The case-based interventions included case detection and isolation through sensitive surveillance systems, contact tracing of confirmed cases to early detect secondary cases among close contacts, and 14-day quarantine of close contacts (regardless of symptoms). The population-based measures included (mostly voluntary) face mask use, personal hygiene, and physical distancing^10,13^.

Despite Taiwan’s initial success, it remains unclear which components among these various interventions contributed significantly to containment. Better understandings of the intervention effectiveness would not only help the preparedness for the next wave but also assist with phased control measures, especially during re-opening^1–3,14^. This post hoc evaluation is particularly crucial as strict border control and drastic physical distancing has considerable short-term and long-term socio-economic repercussions. Several modeling studies examined the effectiveness of case-based or population-based interventions. However, most of them focused on the “what-if” analyses through simulating hypothetical scenarios without empirical linkage to specific real-world settings and primary data^14–18^. Using the epidemiological data and detailed contact tracing information in Taiwan, we applied a stochastic branching model with two-stage calibration to evaluate the effective reproduction number under case-based interventions (*R_c*), population-based interventions (*R_p*), and both (*R_pc*) and to quantify the impact of case-based interventions and population-based measures on the containment of the epidemic.

## Results

### Epidemiology and transmission dynamics of COVID-19 in Taiwan

The epidemic started with few imported cases from China, leading to non-sustained local transmission during January–February 2020 (Fig 1a). In March, a surge of imported cases mainly from North America and Europe caused sporadic local transmission. The capacity of laboratories testing for COVID-19 with reverse transcription-polymerase chain reaction (RT-PCR) was initially low and thus diagnostic testing was limited to symptomatic presumptive cases with relevant travel or contact history (Fig 1a)^11^ The criteria of notification and testing were gradually expanded to include individuals with respiratory or alarming symptoms, with an overall test positive rate of 0.61% by June 1.

**Fig. 1.**
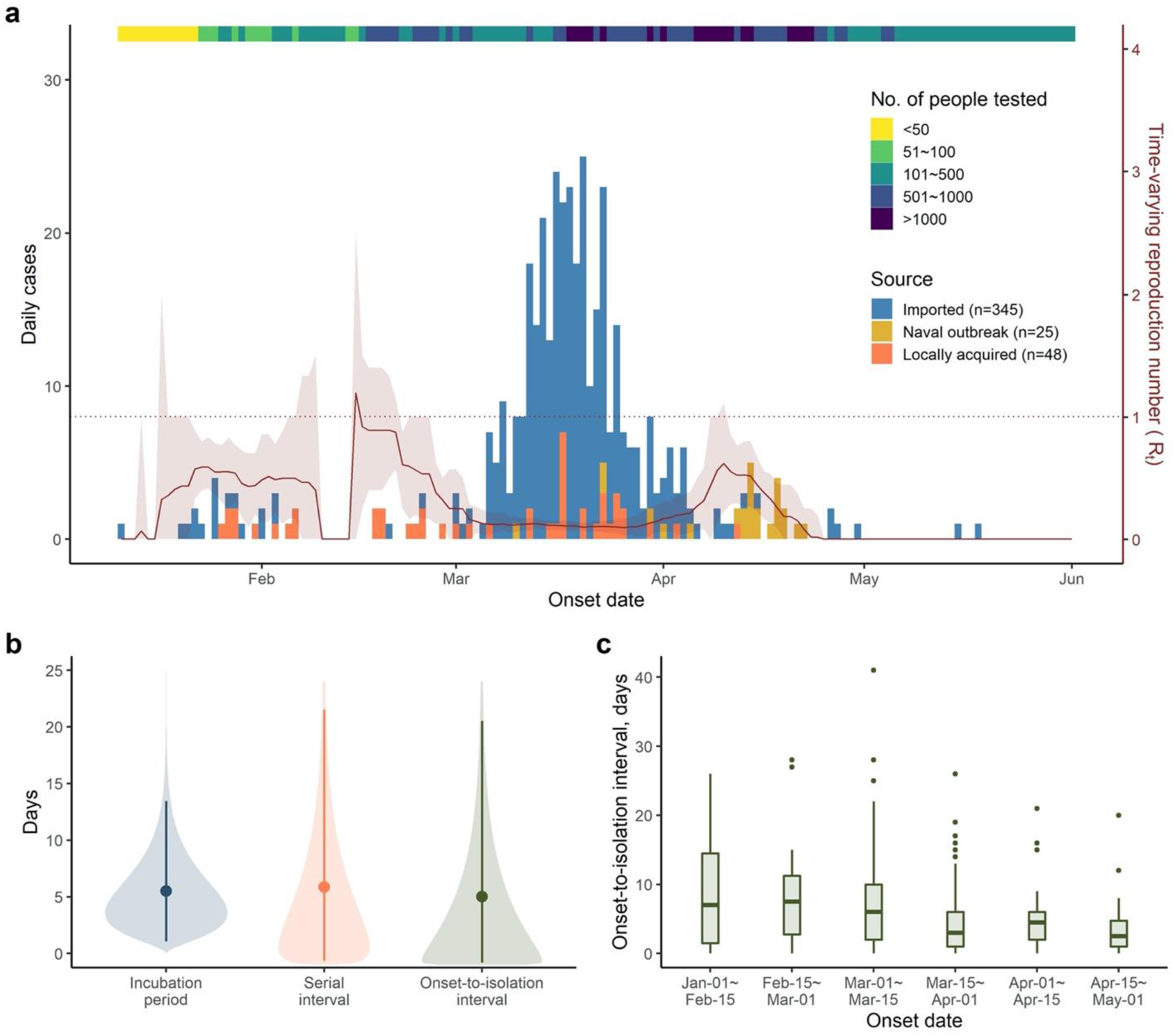
Epidemiological characteristics and parameters of the coronavirus disease 2019 in Taiwan, January 10–June 1. (a) The epidemic curve, number of diagnostic tests, and the time-varying reproduction number (b) Distribution of the incubation period, the serial interval, and the onset-toisolation interval. The points and the intervals represent the mean estimates and the 2.5 and 97.5 percentiles of the estimated distribution. The shaded areas represent the mean estimation of the interval distribution (c) Box-plot of the distribution of the onset-to-isolation interval by onset date. Central lines indicate the median, boxes indicate upper and lower interquartile ranges, whiskers indicate the upper and lower adjacent values (within 1.5-fold the interquartile range), and isolated points indicate outliers.

We analyzed the epidemiological and contact tracing data of 158 confirmed cases to estimate the incubation period, serial interval, and onset-to-isolation interval (Methods)^19^. The estimated mean incubation period and mean serial interval was 5.50 (95% credible interval [CrI]: 1.06–13.45) and 5.86 (95% CrI: −0.64 to 21.51) days, respectively (Fig. 1b). The mean onset-to-isolation interval was 5.02 (95% CrI: −0.81 to 20.51) days, with a decreasing trend over time (Fig. 1c). By fitting the stochastic branching model to the observed serial intervals (Methods), we estimated that 52% (95% CrI: 39–67%) of transmission events occurred during the pre-symptomatic stage.

### Effects of case-based interventions

We used the fitted stochastic branching model and parameter values estimated from the empirical data to simulate the potential impact of case-based interventions (Table 1 for parameter values, Supplementary Notes for details of cased-based interventions). We found that the combination of case detection, contact tracing, and 14-day quarantine of close contacts (regardless of symptoms) could lower the reproduction number under case-based interventions (*R_c*) from the counterfactual value of 2.50 (*R_0_*) to 1.25 (95% CrI: 1.22–1.28) (Fig. 2a). With 100 initial cases introduced to the community (i.e., cases escaping from border control and quarantine), the estimated probability of epidemic extinction was 0% (95% CrI: 0–0%). In the one-way sensitivity analysis, the value of onset-to-isolation interval had the most significant impact on the *R_c*, followed by the incubation period, the proportion of pre-symptomatic transmission, and the counterfactual *R_0_* (Extended Data Fig. 1). Notably, the extinction probability was consistently 0% when the counterfactual *R_0_* was set to 2–3. When the onset-to-isolation interval was shortened to the lowest biweekly value of 2.5 days observed in Taiwan (Fig. 1c), the *R_c* was 1.04 (95% CrI: 1.01–1.07), and the extinction probability was still 0% (95% CrI: 0–0%).

**Table 1.**
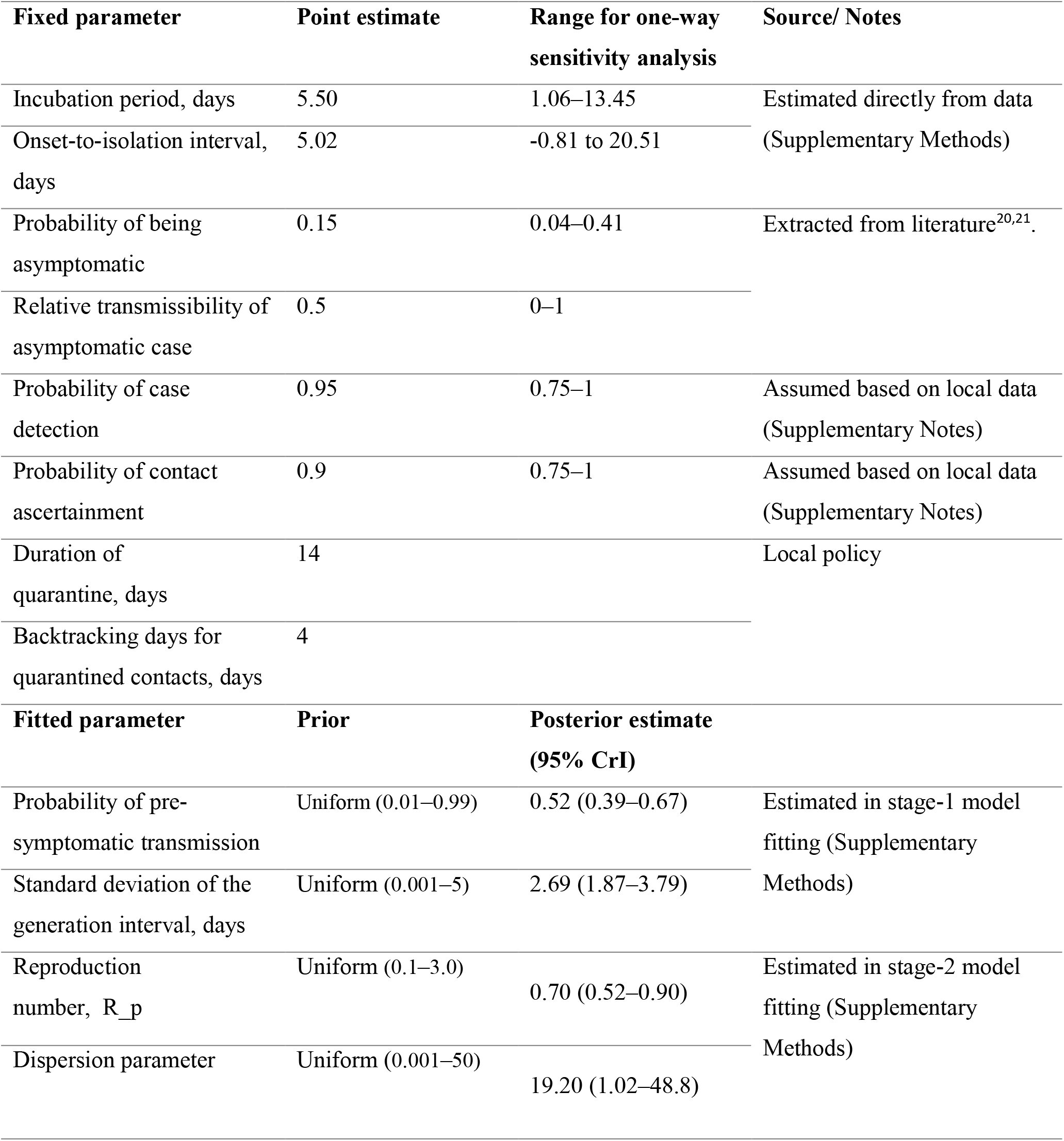
Parameters for the branching process model.

**Fig. 2.**
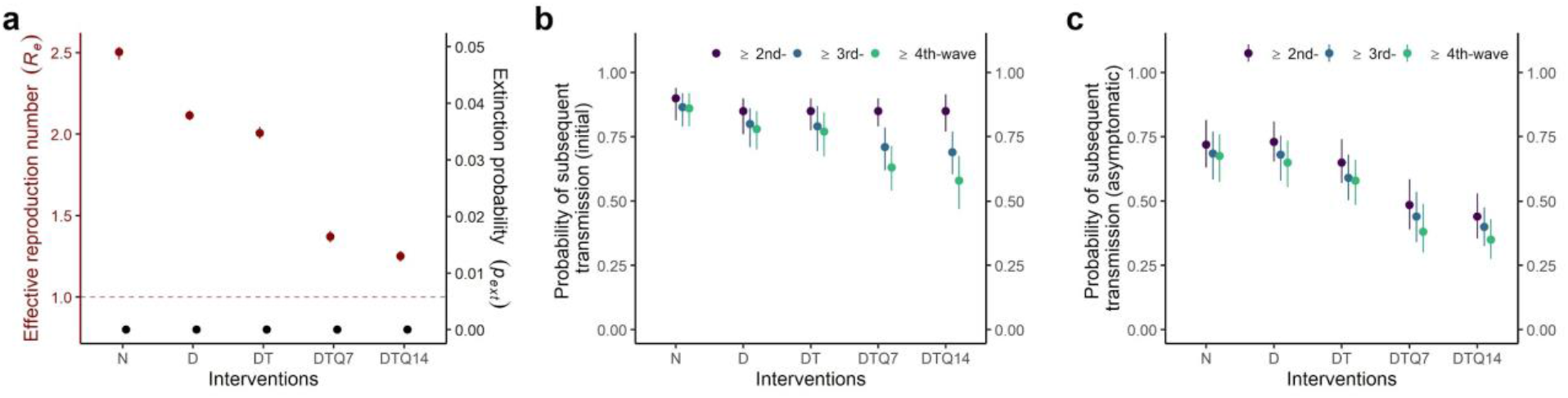
Impact of different combinations of case-based interventions on the effective reproduction number (*R_c*) and transmission of COVID-19 using the fitted stochastic branching model. Results were based on 1000 stochastic simulations under the counterfactual *R_0_* value of 2.50 and 100 introductions. (a) Estimated *R_c* (red dots) and extinction probability (black dots). (b) The probability of subsequent transmission from an index case. (c) The probability of subsequent transmission from any asymptomatic case in the transmission network. N: no case-based intervention; D: case detection; T: contact tracing, Q7/Q14: quarantine of contacts for 7 or 14 days.

Among the different case-based interventions, quarantine of contacts contributed the most to the reduction of *R_c* (Fig. 2a). The case-based interventions had only limited impacts on secondary transmissions, but tertiary and quaternary transmission events could still be reduced (Fig. 2b), and resulted in a lower probability of asymptomatic transmission (Fig. 2c).

### Effects of population-based interventions

Since case-based interventions alone were not sufficient, population-based interventions must also have contributed to the containment. We firstly fitted the model to the observed cluster size distribution to infer the reproduction number under population-based interventions only (*R_p*), assuming a uniform prior distribution (0.10–3.00) of *R_p*. The posterior estimate of *R_p* was 0.70 (95% CrI: 0.52–0.90), suggesting a 62%, 69%, and 75% reduction when the counterfactual *R_0_* was 2.00, 2.50, and 3.00 respectively (Extended Data Fig. 2). The posterior estimate of *R_pc* using the fitted model was 0.49 (95% CrI: 0.41–0.59), which was similar with the *R_pc* directly estimated from the average size of clusters (*R_pc*: 0.36 [95% CrI: 0.14–0.50]). We note that this estimation procedure of *R_p* and *R_pc* was not influenced by the value of counterfactual *R_0_*.

Because the population-based interventions would have collateral impacts on other respiratory infections^6,22^, we quantified the real-time reproduction number (*Rt*) of influenza during the COVID-19 epidemic in Taiwan based on time-series data of influenza cases with severe complications (a notifiable condition in Taiwan), the consultation frequency of influenza-like illness, and the proportion of influenza-positive specimens among the samples of patients with respiratory infection. In this analysis, we found an early and sustained decline of cases in the 2019–20 season compared to the 2017–18 and 2018–19 seasons (Fig. 3a, Extended Data Table 1). The estimated *Rt* of influenza in 2020 dropped from 0.87 on January 21 (when the first case COVID-19 was reported) to 0.20 one month later, corresponding to a 77% decline. Analysis of estimated influenza incidence showed a similar pattern, with a 46% *Rt* reduction from 1.07 on January 21 to 0.58 after January 21 (Fig. 3b).

**Fig. 3.**
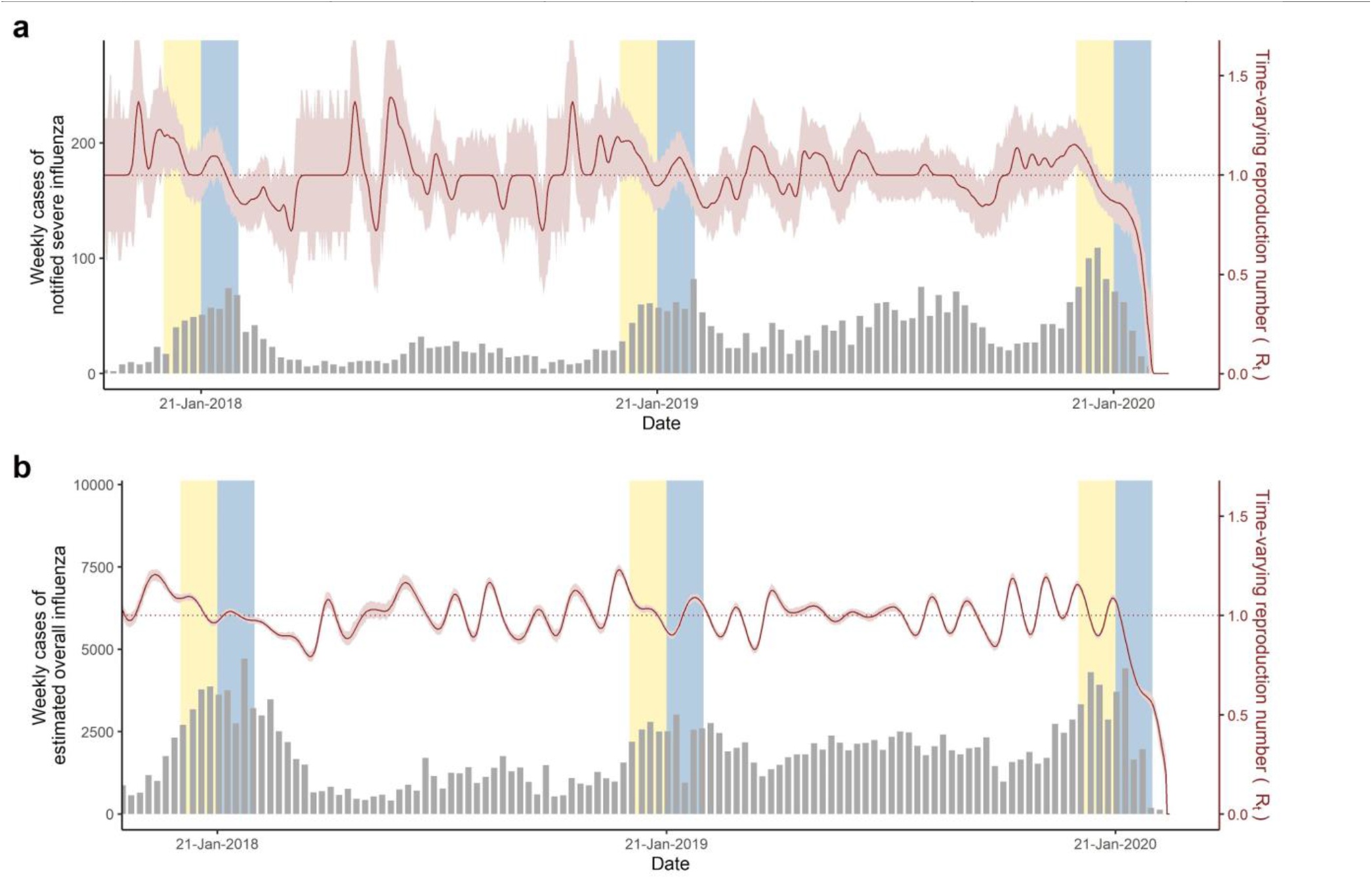
The incidence and real-time reproduction number (*Rt*) of influenza in Taiwan, 2018–2020. (a) Estimates from the notified number of severe influenza with complications. (b) Estimates from the overall influenza cases derived from influenza-like illness consultation rate and the positive rate from laboratory testing for influenza (Supplementary Methods). The gray bars represent the number of weekly incidence cases, and the red lines represent the *R_t_* with 95% confidence intervals in the shaded area. Thirty days before and after (yellow and blue background) January 21, the date of the first SARS-CoV-2 infection confirmed in Taiwan, was highlighted.

### Joint effects of case-based and population-based interventions

We projected the epidemic curve with 100 initial cases under different scenarios using the fitted model (*R_0_*: 2.5, *R_c*: 1.25, *R_p*: 0.70, *R_pc*: 0.49). The case-based interventions could partially suppress the rate of increase, but exponential growth would continue and the daily number of new cases would rise to 5,617 (95% CrI: 3,265–8,335) by day 60 (Fig. 4a). In contrast, we estimated that the effects of population-based interventions would be sufficiently large to control the epidemic and lead to local extinction, provided that behavioral changes could be maintained. Combining case-based and population-based interventions would more quickly control the epidemic and provides insurance in case of lapses. We further estimated the probability of successful COVID-19 containment in Taiwan with case-based interventions under different values of *R_p* and initial numbers of introductions (representing variation in the prevalence of infection in travelers and the effectiveness of border controls with corresponding quarantine requirement) (Fig. 4b). When the number of introductions was set to greater than 100, *R_p* had to be kept below 1.6 to achieve an extinction probability of at least 90%. If *R_p* was greater 2.5, it would be impossible to rely only on case-based interventions to contain the outbreak (extinction probability: 0) even when the number of introductions was small.

**Fig. 4.**
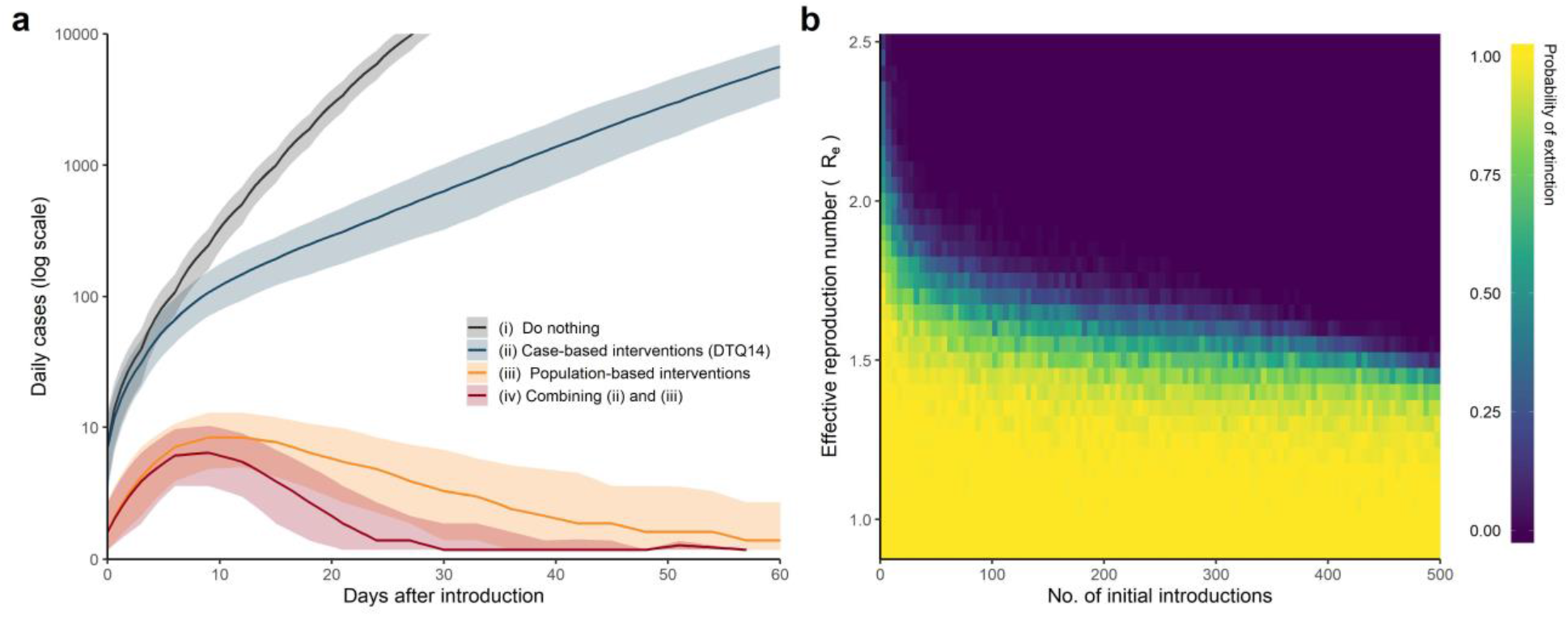
Projections and joint effects of case-based and population-based interventions on probability of epidemic extinction. (a) The projected epidemic curves with 100 initial introductions under different scenarios regarding the intervention being implemented. The four scenarios included: (i) no intervention, (ii) case-based interventions only, (iii) population-based interventions only, and (iv) combining both case-based and population-based interventions. We assumed a sensitivity of 95% for case detection, an ascertainment probability for contact tracing of 90%, and a 72% reduction in background *R_0_* by population-based interventions (*R_0_*: 2.5, *R_p*: 0.7). The uncertainty intervals were calculated by the 2.5th and 97.5th percentiles from 1000 replicate simulations. (b) The probability of epidemic extinction using case-based interventions (detection, contact tracing, and 14-day quarantine of close contacts) under different levels of population-based interventions (*R_p*) and initial numbers of introductions. Each cell presents the estimated probability of extinction based on 100 replicate simulations using the stochastic branching model.

### Discussion

Using a flexible modeling approach that incorporated multiple sources of primary data, we estimated and compared the impact of case-based and population-based interventions in Taiwan. We found that case-based interventions alone were not sufficient to contain the epidemic, even in a setting where the public health and healthcare system was not yet overwhelmed, and a highly efficient contact tracing program was in place. We also found that the mostly voluntary population-based interventions reduced the reproduction numbers by more than 60% and played a crucial role in the containment in Taiwan. Our analysis of Taiwan’s success highlights that coordinated efforts from both the government and the citizens are indispensable in the fight against COVID-19 pandemic.

Previous modeling studies showed promising effectiveness of contact tracing with corresponding management of contacts (either quarantine or active monitoring) in settings with well-functioning public health programs like Taiwan^15–17^. However, our results suggested that even in a well-prepared setting, contact tracing alone would fail to eliminate an epidemic when multiple introductions were likely. This discrepancy is driven primarily by the growing understanding of the role of pre-symptomatic transmission and the challenge this presents for shortening the delay from symptom onset to isolation. The effectiveness of contact tracing depends on the timeliness of case detection and implementing corresponding quarantine for high-risk contacts^16^. When the timeliness of case-based interventions has been optimized, the remaining transmissions can be blocked only by implementing more generalized non-symptom-based approaches. According to our analysis of local data, the mean onset-to-isolation interval was around five days in Taiwan. This relatively long delay compared to the short serial interval suggests the virus usually had already been transmitted in the community by the time locally-acquired infections were detected. Therefore, population-based interventions should be prioritized over case-based interventions, even in settings where case-based interventions are feasible. That said, we did find case-based interventions to have significant impact on their own, and they provide additional assurance that the epidemic can be controlled and brought to extinction more quickly and reliably than under use of only population-based interventions.

On the other hands, we found that population-based interventions played a major role in containment. A meta-analysis of 172 observational studies in healthcare and non-healthcare settings revealed that physical distancing, face mask use, and eye protection were significantly associated with reduced COVID-19 transmission at the individual level^23^. The population-level effects of these interventions were assessed by other scenario-based analyses without empirical evaluation^3,15–17,24^ Our study is among the very few that directly and empirically evaluated the population-level impact. In Taiwan, several essential practices have been in place after the 2003 SARS outbreak and 2009 H1N1 pandemic. For example, face mask use was common among people with respiratory symptoms, and school children were advised to avoid attendance when fever developed. Protocols of class/school closure in response to an outbreak were also developed and implemented. These existing practices may contribute to the resilience of the society against respiratory infection outbreaks^22^.

This study provides crucial insights into the role of different control measures at different stages and settings. Border control with corresponding quarantine for entering travelers may be an option to limit the epidemic in the beginning by reducing the number of introductions, especially in certain contexts (e.g., island nations like Taiwan and New Zealand)^8^. In Taiwan, the quarantine requirement for travelers returning from other countries after March 21 effectively limited the number of introductions in the community. However, the effect of border control with quarantine would be quickly diminished when the number of introductions exceeds 100 (Fig. 4b), and this has important implications for the possibility of new epidemics in places like Taiwan that have achieved successful control over the first wave. This finding might also explain the resurgences that occurred in Hong Kong, Vietnam, and New Zealand when travel restrictions were lifted after a period of epidemic^25,26^. With a sufficiently large number of introductions, the probability of local transmission is high, and containment efforts should shift toward suppressing local transmission using both case-based and population-based interventions. Our analysis revealed that blended approaches could reliably result in epidemic extinction, even when the number of introductions is high. When discussions on re-opening borders to boost economic growth raise, our results suggest that travel restrictions may be judiciously lifted only when other effective interventions can be maintained. The stringency of border control can be calibrated by pre- or post-arrival testing and entry quarantine, depending on the infrastructure capacity of individual countries and the number of COVID-19 cases that can be managed by local public health and healthcare system.

We note an interdependency between case-based and population-based interventions^15^. For example, contact tracing and quarantine would be increasingly difficult in practice when the number of cases surges. In this case, the intensity of population-based interventions has to be strengthened to compensate for the decreased efficiency of case-based interventions. On the other hand, if the population-based interventions can effectively suppress local transmission, it would require less effort from the public health workforce to contain the community outbreak. Nonetheless, maintaining behavioral changes and adopting the new normal is challenging in many places^27^. Since the health behaviors may change rapidly over time, monitoring these behaviors in the community would be informative^6^. In our analysis, we explored the potential of using influenza activity as a proxy indicator to monitor the *Rt*^28^. Continuous measurement of the implementation and impacts of case-based and population-based interventions would help to calibrate the efforts in contact tracing and quarantine and to reserve the precious public health resources.

One limitation of this study is that we could not directly estimate the “counterfactual” *R_0_* in the absence of any interventions since the majority of case-based and population-based interventions were triggered soon after the outbreak begun. Nonetheless, our main conclusions were robust within the range of commonly reported *R_0_* values of 2–3. Additionally, the analysis of cluster size distribution relied on the assumption of complete information of all clusters. If small clusters were more likely to be missed by surveillance and contact tracing, the estimated reproduction number (*R_pc*) would have been lower, and the impact of population-based intervention would have been larger.

### Conclusion

Through the analysis of a presumably high-risk country, we found that the combination of case-based interventions from an effective public health system and voluntary behavioral changes in a vigilant population likely contributed to the initial success of epidemic control. Notably, case-based interventions alone may not be sufficient to contain COVID-19, given the significant role of pre-symptomatic transmission. Population-based interventions were the key to success in our setting. Still, the longer-term sustainability and generalizability of these behaviors (e.g., face mask use) to other settings must be assessed.

## Methods

### Data

Case series data of SARS-CoV-2 infections in Taiwan were collected from the official website of Taiwan Centers for Disease Control (TCDC) and reviewed by TCDC officers to clarify missing information. All the cases were confirmed by RT-PCR tests using one of the following clinical specimens: nasopharyngeal swab, throat swab, expectorated sputum, or lower respiratory tract aspirates^29^. Cases were isolated immediately after being notified to Taiwan CDC.

We analyzed the epidemiological and contact tracing data to capture the transmission dynamics of COVID-19 in Taiwan^19^. Starting March 21 all inbound passengers (citizens and eligible non-citizens) to Taiwan were required to undergo a 14-day quarantine upon entry, and nearly all confirmed cases afterward were imported and were diagnosed during the quarantine period (Supplementary Notes). We, therefore, enrolled locally-acquired cases, epidemiologically-confirmed clusters, and imported cases who entered Taiwan before March 21 and excluded the returnees who were tested at the airport or diagnosed during home quarantine (final sample size: 158 cases with 3154 contacts).

### The stochastic branching process model

We adapted the stochastic branching process model developed by Hellewell, et al^17^. The model generated transmission trees by drawing the number of secondary cases, based on the reproduction number and the dispersion parameter. Transmission either occurred or prevented, depending on the comparison between the sampled generation interval and the isolation or quarantine period of the index case (Extended Data Fig. 4). We parameterized the distribution of generation interval as a skewed normal distribution centered at each index case’s onset time to avoid discordant incubation period and generation interval. The shape of generation interval distribution was determined by the proportion of pre-symptomatic transmission and the standard deviation of generation interval. Distributions of the incubation period, serial interval, and onset-to-isolation interval were independently estimated from data using Bayesian hierarchical models (as described in Supplementary Method 1, Supplementary Table 1 for details of the branching model and parameterization).

### Parameter estimation and model fitting

We estimated the unknown key parameters by fitting the branching model to empirical data using two-stage calibration with the sequential Monte Carlo algorithm^15,30^. In the first stage, the model was fitted to the observed serial intervals to estimate the proportion of pre-symptomatic transmission and the standard deviation of generation interval. In the second stage, the model was fitted to the cluster size distribution (for all epidemiologically-linked clusters) to estimate the reproduction number under population-based measures only (*R_p*) (see below). A wide uniform distribution was assumed as the prior for each estimated parameter (Supplementary Method 2, Supplementary Table 1–2).

### Estimating the effects of case-based and population-based interventions

We estimated the effect of case-based interventions using the stochastic branching model after stage-1 fitting. Since the majority of these containment measures were in place at the beginning of the epidemic, it was not possible to directly estimate the “counterfactual” *R_0_* (the hypothetical reproduction number without interventions) in Taiwan. We assumed this counterfactual *R_0_* to be 2.50 (range 2–3), similar to the estimated *R_0_* in Hong Kong at the beginning of its outbreak and consistent with the previously estimated *R_0_* values^6,31,32^. Five scenarios were considered: (1) no case-based interventions; (2) case detection and isolation; (3) case detection and contact tracing to detect and isolate secondary cases; (4) case detection, contact tracing, and 7-day quarantine for contacts (regardless of symptoms), (5) case detection, contact tracing, and 14-day quarantine (Supplementary Table 3). Whereas the 14-day quarantine for contacts was a current policy implemented in Taiwan, we simulated the scenario with 7-day quarantine to see the trend of impact of quarantine duration. The primary indicator was the mean effective reproduction number, along with the probability of outbreak extinction, which was defined as zero new cases within 20 generations.

The effect of population-based interventions was estimated using two independent approaches. First, we analyzed the observed cluster size distribution in Taiwan to estimate the effective reproduction number, which was affected by both case-based and population-based interventions (hence the estimated reproduction number should be *R_pc*). First, since the impact of case-based interventions was explicitly incorporated in the branching process model, we could estimate the reproduction number under population-based measures only (*R_p*) by fitting the dynamical model to the observed cluster size distribution (Supplementary Method 3). The posterior estimation of *R_pc* was also compared to the analytic estimation using the average size of clusters (*R* = 1 − 2/m, where m is the expected average size for clusters with ≥ 2 cases)^33^. Second, we estimated the time-varying reproductive numbers (*Rt*) of seasonal influenza and used this as a proxy measure of the impact of population-based interventions on control of respiratory infections (Supplementary Method 4). The *Rt* of influenza was estimated using the time-series data of influenza cases with severe complications (a notifiable condition in Taiwan), the consultation frequency of influenza-like illness, and the proportion of influenza-positive specimens among the samples of patients with respiratory infection. These data were obtained from influenza surveillance systems of Taiwan CDC^34,35^.

## Data Availability

The datasets generated and analyzed during the current study are available from the corresponding author on reasonable request.

## Code availability

Codes are available on GitHub at https://github.com/dachuwu/Taiwan_CovidDTQ before publication.

## Author contributions

T.C.N., H.Y.C., and H.H.L. conceived and designed the study. T.C.N., H.Y.C., C.C.L., C.C.Y, and S.W.J collected data. T.C.N., C.C.L., and C.C.Y performed the data analysis. T.C.N., H.H.C, and H.H.L. built the model. H.Y.C, H.H.C, D.P.L., T. C., and H.H.L. interpreted the findings. H.H.L supervised the study. T.C.N, H.Y.C, and H.H.L drafted the manuscript and all authors contributed to the revision of the final manuscript.

---

**Extended Data Fig. 1.**
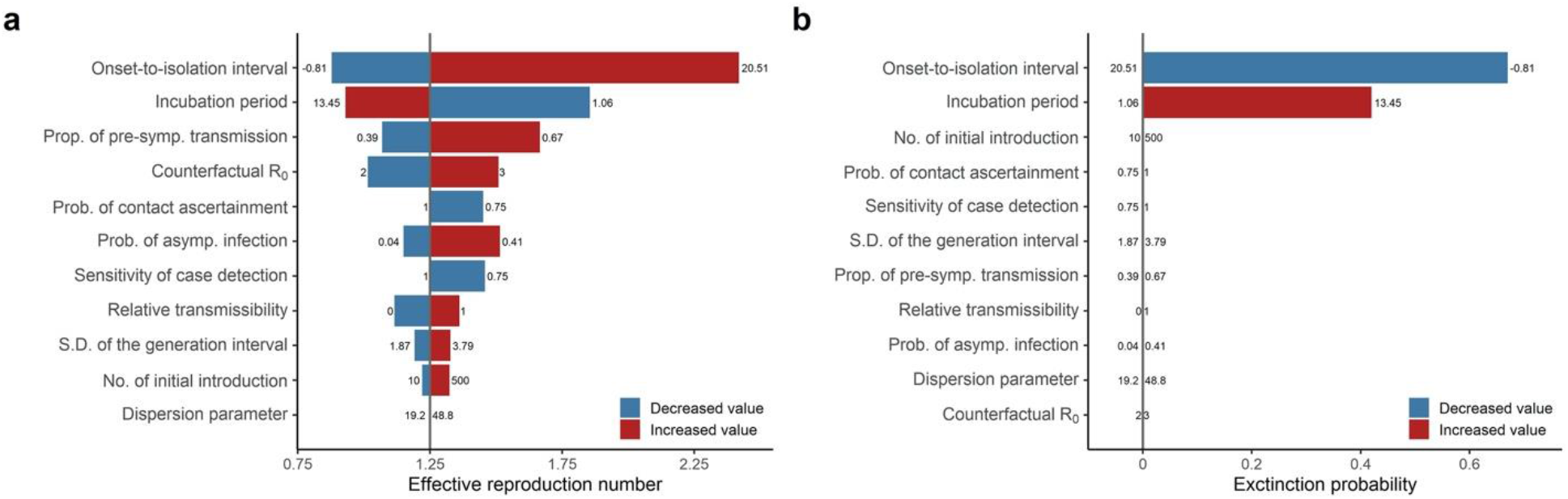
Tornado diagrams from the one-way sensitivity analysis on the effects of case-based interventions. (a) The effective reproduction number under case-based interventions. (b) The extinction probability. The blue bars represent the change in the measured outcome when the corresponding parameter value decreased; the red bars represent the change when the parameter value increased. The tuning ranges of the parameters are shown next to the bars.

**Extended Data Fig. 2.**
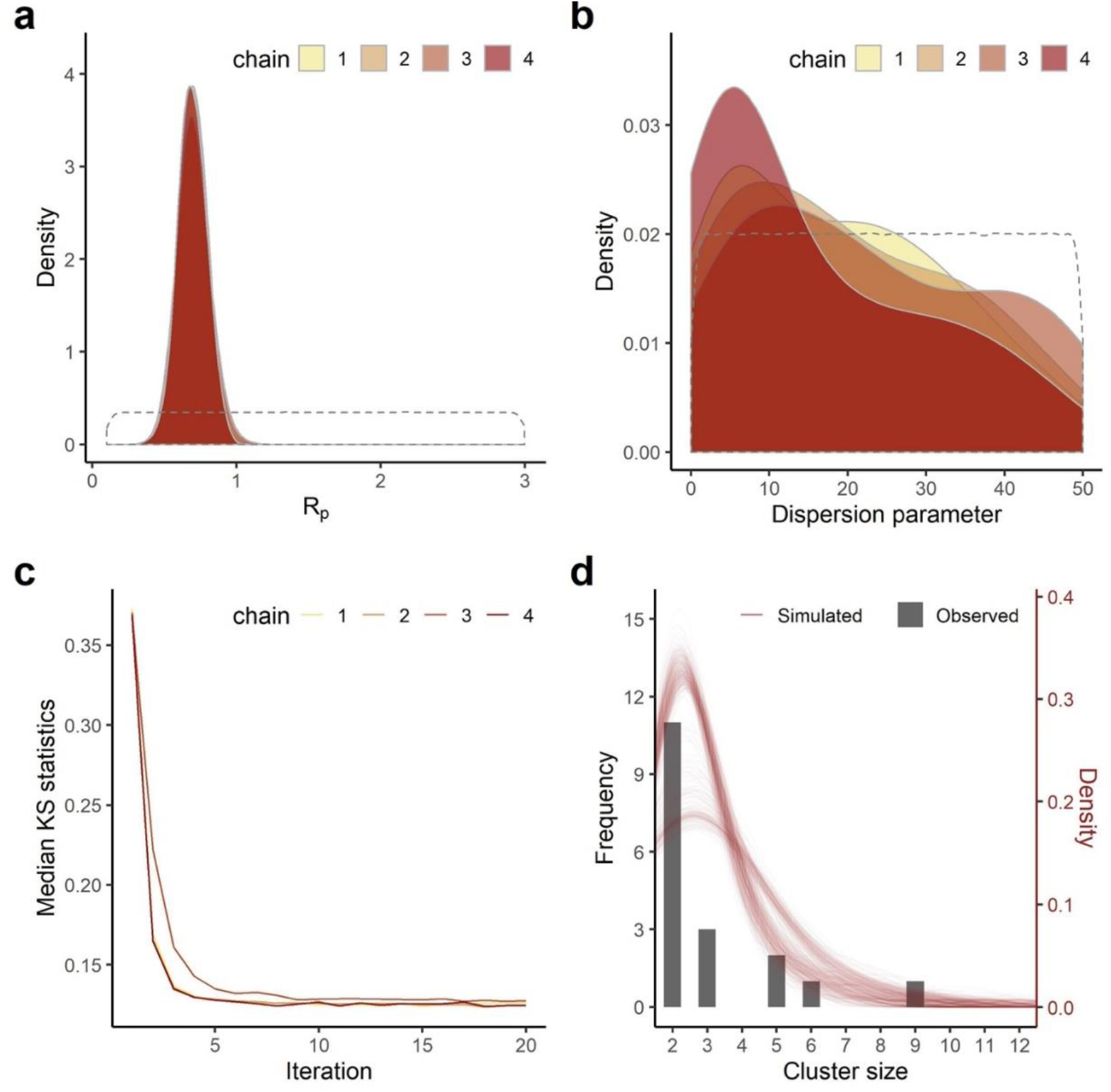
Model fitting to the observed cluster sizes. (a) The posterior distribution of the effective reproduction number under population-based interventions (*R_p*). (b) The posterior distribution of the dispersion parameter. (c) The convergence plot of the sequential Monte Carlo algorithm, with the median Kolmogorov–Smirnov (KS) statistics as the distance measure. (d) The observed cluster sizes and the simulated cluster size distribution by the fitted model (100 times of simulation).

**Extended Data Fig. 4.**
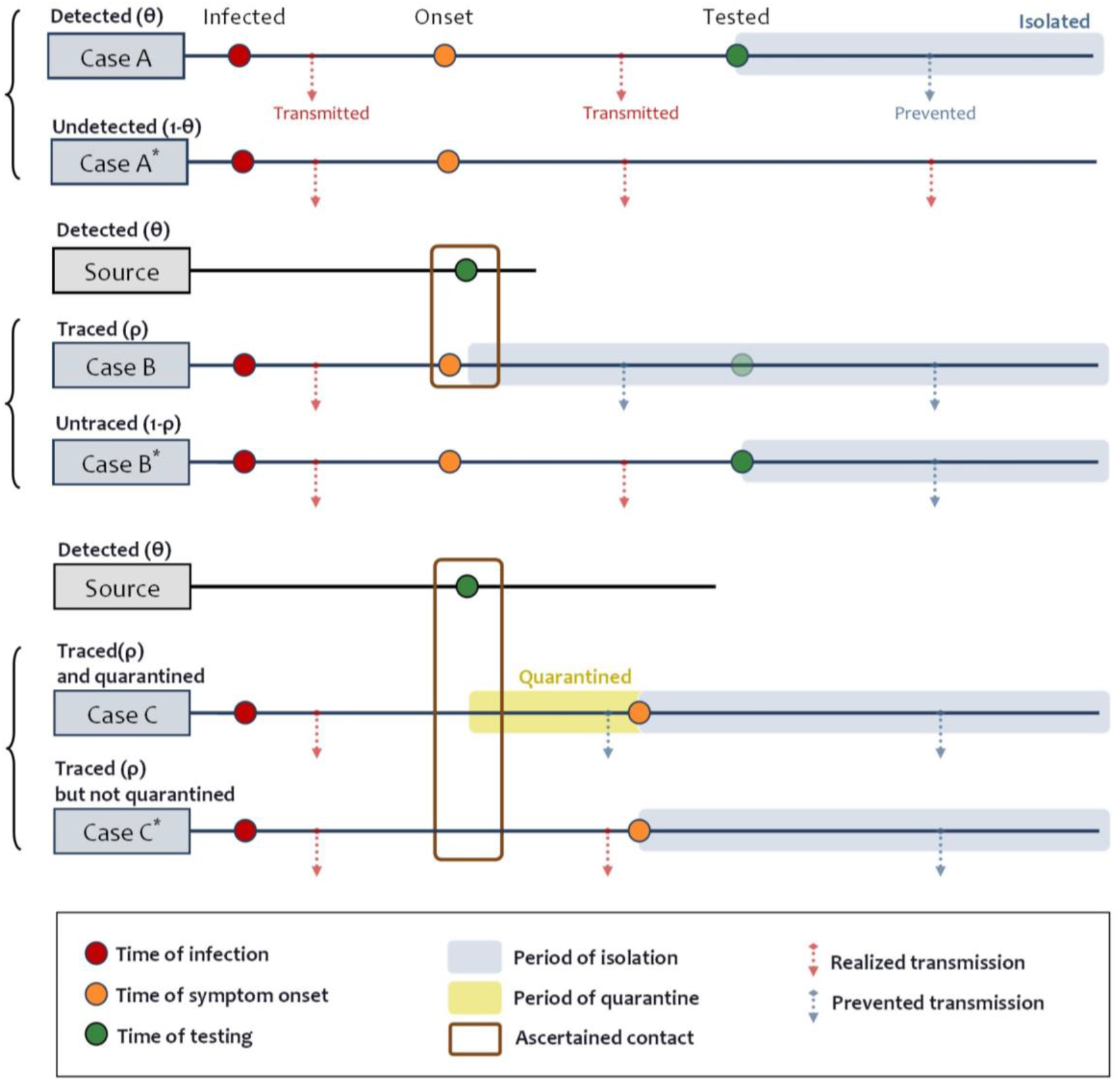
Examples of the effects of case detection, contact tracing, and quarantine. Case A and A* demonstrate the effect of mere detection, which can only prevent the transmission once the active cases are tested and isolated. That is, the active cases can transmit the disease during their incubation and delay of case detection. Case B and B* demonstrate the effect of detection plus tracing (without quarantine), where case B was successfully traced and onset within a buffer period. Therefore, case B was immediately isolated when the source was detected. Detection plus tracing can prevent transmission during the delay of case detection. Case C and C* demonstrate the combined effect of detection, contact tracing, and quarantine. Only in this scenario that transmission during the incubation period can be prevented. We assume that there is no delay between testing and isolation, the buffer of contact tracing to be one day, and the same effect of quarantine and isolation. Besides, asymptomatic cases are never detected or traced but could be quarantined and have lower transmissibility.

**Extended Data Table 1.** The estimated real-time reproduction number (*Rt*) of influenza in Taiwan on January 21, February 4, and February 18, 2018–2020. The daily incidence of influenza was derived from the notified influenza patients with severe complications or an estimated overall number of medical visits cases due to influenza-like illness.

| Year | Notified severe influenza |  |  | Estimated total influenza |  |  |
| --- | --- | --- | --- | --- | --- | --- |
|  | January 21 | February 4 | February 18 | January 21 | February 4 | February 18 |
| 2018 | 1.01<br>(0.92–1.16) | 1.09<br>(0.95–1.24) | 0.89<br>(0.77, 0.99) | 0.97<br>(0.96–0.99) | 1.01<br>(1–1.03) | 0.98<br>(0.96–1) |
| 2019 | 0.95<br>(0.82–1.08) | 1.07<br>(0.91–1.18) | 0.96<br>(0.84, 1.06) | 0.92<br>(0.91–0.94) | 1.01<br>(0.99–1.04) | 1.07<br>(1.05–1.09) |
| 2020 | 0.87<br>(0.77–0.98) | 0.79<br>(0.65–0.92) | 0.2<br>(0, 0.43) | 1.07<br>(1.06–1.09) | 0.72<br>(0.71–0.74) | 0.58<br>(0.54–0.61) |

